# Evidence linking atopy and staphylococcal superantigens to the pathogenesis of lymphomatoid papulosis, a recurrent CD30+ cutaneous lymphoproliferative disorder

**DOI:** 10.1101/19012088

**Authors:** Marshall E. Kadin, Robert G. Hamilton, Eric C. Vonderheid

**Affiliations:** Boston University and Research Scientist, Lifespan-Rhode Island Hospital, 593 Eddy Street. Providence RI; Johns Hopkins University School of Medicine and Director Johns Hopkins Dermatology, Allergy and Clinical Immunology Reference Laboratory; Sydney Kimmel Cancer Center, Johns Hopkins University School of Medicine

**Author notes:** The authors declare no conflicts of interest.

**Keywords:** Atopy, IgE, cutaneous lymphoma, lymphomatoid papulosis, Staphylococcus, superantigens, enterotoxins

## Abstract

**Background:** Lymphomatoid papulosis (LyP) is a self-healing CD30+ cutaneous lymphoproliferative disorder (CLPD) with paradoxical histology of a malignant lymphoma. Case reports and small patient series suggest an association of LyP with atopy. However, the prevalence of atopy depends on patients’ recall which is not always reliable. More objective criteria of atopy include skin reactivity to allergens and IgE reactivity to allergens. This study was undertaken to determine if atopy is prevalent in LyP patients using IgE specific antibodies to aeroallergens, and if Staphylococcal aureus enterotoxins might be a pathogenic factors for LyP as proposed for other skin disorders.

**Methods:** Thirty-one samples of CD30CLPD were tested for total serum IgE (IgE-t) and 10 IgE-specific airborne allergens with the Phadiatop multiallergen test, which if positive, is regarded as evidence of atopy. Sera was tested for IgE reactive against three Staphylococcal enterotoxins with superantigenic properties (SSAg-IgE). Control sera were obtained from adult subjects evaluated for rhino-sinusitis and a negative Phadiatop test. Patients’ history of atopy was obtained by retrospective chart review.

**Findings:** Nearly 50% of patients with the most common LyP types had a positive Phadiatop test and IgE-t was increased compared to non-atopic controls. Seven of 28 (25%) LyP patients had at least one SSAg-IgE at the concentration used to define serologic atopy (≥ 0.35 kUa/L) compared to 3/52 (6%) controls (P= 0.028). TSST1-IgE was detected in 7 (23%) specimens of CD30CLPD, often together with SEB-IgE; SEA-IgE was not detected. TSST1-IgE exceeded the 0.35 kUa/L threshold in 3 (6%) controls.

**Conclusions:** LyP patients have an increased prevalence of atopy as determined by the Phadiatop test and increased prevalence of SSAg-IgE compared to controls. Prevalence of serologic atopy exceeded that reported by patients’ medical history. The results support the hypothesis that an atopic diathesis and possibly SSAg contribute to the pathogenesis of LyP.

**Summary:** Contrary to patients’ recall of atopic disorders, IgE specific antibodies to aeroallergens, Staphylococcal aureus superantigens and total serum IgE are increased in patients with lymphomatoid papulosis.

## Introduction

Primary cutaneous CD30+ lymphoproliferative disorders (CD30CLPD) are the second most common type of cutaneous T cell lymphoma (CTCL) and include lymphomatoid papulosis (LyP) and primary cutaneous anaplastic large cell lymphoma (pcALCL) at benign and malignant ends of the spectrum, respectively.(1) LyP is characterized clinically by spontaneously regressing papules and nodules (usually less than 2 cm diameter) and is divided into five subtypes A to E based on histo-immunopathologic findings.

LyP-A, the most common expression of LyP, is characterized by scattered large CD30+ CD4+ cells in the dermal infiltrate together with lymphocytes and other inflammatory cells (neutrophils and eosinophils). LyP-B has histopathologic features resembling mycosis fungoides with atypical small-intermediate sized lymphocytes with cerebriform nuclei that usually do not express CD30. LyP-C has a dermal infiltrate with large clusters or sheets of atypical CD4+CD30+ cells that might occur in pcALCL.The two remaining subtypes of LyP are CD8+ rather than the more typical CD4+ variants. LyP-D is characterized by a dense pagetoid infiltrate of the epidermis as well as dermis by atypical cells that express a CD3+CD4-CD8+ phenotype (rarely CD3+CD4-CD8- phenotype) and CD30 to a variable degree. LyP-E is an extremely rare angiocentric variant with CD8+CD30+ cells. pcALCL is defined clinically by large nodules, plaques or tumors that tend to persist although spontaneous regression can occur in up to 40% of lesions. The dermal infiltrate typically contains sheets of large CD30+ cells with or without other inflammatory cells. However, some cases of clinical pcALCL have a dermal infiltrate more typical of LyP-A. Such cases have been designated grade III pcALCL to distinguish them from more typical cases of pcALCL (grade IV).(2) Thus, there is an overlap between LyP-A and grade III pcALCL and LyP-C and grade IV pc ALCL.

In a previous study, we reported that total serum Ig-E (IgE-t) levels of patients diagnosed with CD30CLPD are increased compared to published series of non-atopic patients.(3) Moreover, 25/84 (30%) patients with LyP-A, 9/26 (35%) of patients with LyP-C and 5/14 (36%) patients with pcALCL had IgE-t levels greater than 100 kU/L, a threshold used to signify “probable atopy” in adults.(3) None of 5 patients with LyP-B exceeded this threshold and only single instances of LyP-D and LyP-E were encountered. However, IgE-t levels did not correlate with the patients’ personal or family history of an atopic condition. This observation suggested that factors other than genetic predisposition for atopy (atopic diathesis) of patients mediated the increase in IgE-t, and we hypothesized that atypical CD30+ cells might be directly promoting IgE production, possibly by secretion of IgE-enhancing cytokines such as IL-13. (4, 5) Although CD30+ cells in LyP often lack expression of the T cell receptor (TCR) (6) CD30 signaling in the absence of the TCR selectively induces IL-13 production. (5) IgE-t levels may be influenced by other factors including age (IgE levels in pre-adults > adults), gender (IgE levels in men > women), race (IgE levels in non-White > White), use of tobacco (IgE levels in smokers > non-smokers), socioeconomic status, and disease-related factors including bacterial production of superantigens (SAg).(7)

In this study, we re-evaluate the role of atopy and its association with IgE-t levels by screening available serum samples of CD30CLPD for common IgE-specific airborne allergens (aero-IgE) with the Phadiatop multi-allergen screening test, which if positive, is regarded as serologic evidence of atopy.(8) We also test sera for IgE reactive against three Staphylococcal enterotoxins with superantigenic properties (SSAg-IgE) to investigate whether SSAg have a pathogenic role in LyP as has been shown for eruptive guttate psoriasis and Sézary syndrome. Our principal findings are that patients with LyP types A and C have a significant increase in serologic evidence of atopy compared to controls and that the prevalence of atopy based on the Phadiatop test was significantly higher than the prevalence of atopic condtions recorded in the patients’ medical history. In addition, at least one SSAg-IgE was detected at the concentration used to define serologic atopy (≥ 0.35 kUa/L) for 7/28 (25%) patients with LyP which was significantly higher than controls. The serologic findings support the hypothesis that atopy and perhaps SSAg contributes to the pathogenesis of LyP types A and C.

## Methods

Clinical data and previously obtained IgE-t measurements from other reference laboratories were obtained from a Cutaneous Lymphoma Registry approved by the Institutional Review Board at Johns Hopkins University. The LyP study population consisted of 19 patients with LyP-A (7 men, 12 women; median age, 56 years, range 13 to 77 years) and 9 patients with LyP-C (5 men, 4 women; median age, 47 years, range 15 to 80 years). Three samples of pcALCL were also studied (3 men, median age, 54 years, range 19 to 62 years). All but 2 patients were White. The percentage of CD30+ cells in the dermal infiltrate of the skin specimen obtained at the time of evaluation was visually estimated and categorized into 4 groups: < 5%, 5-19%, 20-49% and ≥ 50% CD30+ cells.

Measurements of IgE-t and allergen-specific IgE on available de-identified frozen sera collected at the time of clinical evaluation were obtained in the Johns Hopkins Dermatology Allergy and Clinical Immunology Reference Laboratory using the ImmunoCAP method (ImmunoCAP, Thermofisher Scientific, Uppsala, Sweden). The Phadiatop multiallergen test screens for IgE reactivity against 10 common aero-IgEs in the Northeastern United States. IgEs-specific for Staphylococcal enterotoxin superantigens A,and B (SEA, SEB) and toxic shock syndrome toxin-1 (TSST1) were also measured separately. The lower limit of detection was 2.0 kU/L for IgE-t and 0.1 kUa/L for allergen-specific IgEs. A positive test for aero-IgE or SSAg-IgE was defined as a concentration of ≥ 0.35 kUa/L. Sera collected from adult patients (22 men, 30 women; ages, 24 to 78 years), who were evaluated for rhino-sinusitis and had a negative Phadiatop test, served as a serologic non-atopic control group. Measurement of soluble CD30 (sCD30) in some CD30CLPD patients was performed at ARUP laboratories (Salt Lake City, UT) as part of a previously reported study of cytokine expression.(9)

### Statistics

Results of IgE measurements are given as median value with a range. Because IgE-t and allergen-specific IgE values tend to be skewed, (3, 7) the distribution of these variables was normalized by log transformation and the geometric mean (GM), i.e., the anti-log of the mean log10 IgE-t value, and its 95% confidence interval are also provided. The nonparametric Kruskal– Wallis ranks test was used to test differences of median IgE values among independent groups. In addition, log transformed IgE values were used to compare mean values using parametric tests (t-test with equal variances not assumed, one way analysis of variance and Dunnett’s t-test for comparison against the control sera). Fisher’s and Pearson’s chi-square exact tests were used to test categorical data in 2 by 2 and R by C tables, respectively. Spearman’s rank correlation coefficient was used to determine the strength of association between IgE-t and sCD30 values. The statistical software used in the study were SYSTAT10 and SPSS 13.0 for Windows, SPSS, Inc. (Chicago, IL) and StatXact-3, Cytel, Inc. (Cambridge, MA).

## Results

### IgE-t is increased in patients with LyP

Table 1 summarizes the results of IgE-t according to diagnostic categories in this study. In agreement with our previous study, (3) IgE-t values for LyP-A and LyP-C subgroups were significantly higher than non-atopic controls. IgE-t was not increased for pcALCL, but this was based on only 3 patients. To clarify this issue, IgE-t values from reference labs were analyzed on 12 additional patients with pcALCL not included in this study. The IgE-t GM was 43.6 kU/L (CI, 17.5-108 kU/L). Although lower than the GM of 61.9 kU/L (CI, 23.0-167 kU/L) for patients with LyP-C, the difference was not statistically significant (KW, P= 0.685, t-test, P= 0.580).

**Table 1.**
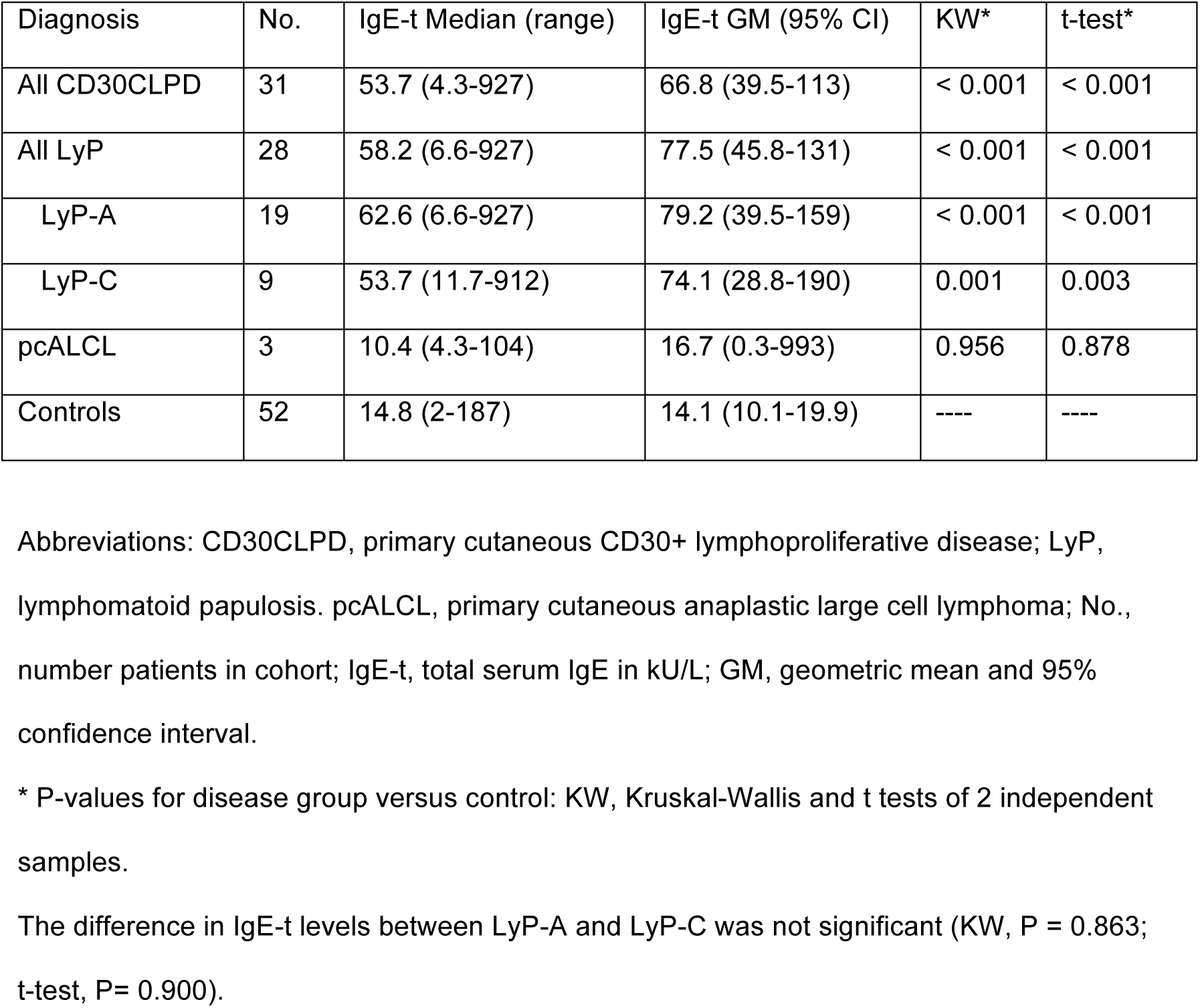
Total serum IgE according to diagnostic categories.

| Diagnosis | No. | IgE-t Median (range) | IgE-t GM (95% CI) | KW* | t-test* |
| --- | --- | --- | --- | --- | --- |
| All CD30CLPD | 31 | 53.7 (4.3-927) | 66.8 (39.5-113) | < 0.001 | < 0.001 |
| All LyP | 28 | 58.2 (6.6-927) | 77.5 (45.8-131) | < 0.001 | < 0.001 |
| LyP-A | 19 | 62.6 (6.6-927) | 79.2 (39.5-159) | < 0.001 | < 0.001 |
| LyP-C | 9 | 53.7 (11.7-912) | 74.1 (28.8-190) | 0.001 | 0.003 |
| pcALCL | 3 | 10.4 (4.3-104) | 16.7 (0.3-993) | 0.956 | 0.878 |
| Controls | 52 | 14.8 (2-187) | 14.1 (10.1-19.9) | ---- | ---- |

Abbreviations: CD30CLPD, primary cutaneous CD30+ lymphoproliferative disease; LyP, lymphomatoid papulosis. pcALCL, primary cutaneous anaplastic large cell lymphoma; No., number patients in cohort; IgE-t, total serum IgE in kU/L; GM, geometric mean and 95% confidence interval.
\* P-values for disease group versus control: KW, Kruskal-Wallis and t tests of 2 independent samples.
The difference in IgE-t levels between LyP-A and LyP-C was not significant (KW, $P = 0.863$ ; t-test, $P = 0.900$ ).

### Clinical atopy and aero-IgE screen (Phadiatop test)

Fifteen of 31 (48%) patients with CD30CLPD had a positive (≥ 0.35 kUa/L) Phadiatop multi-allergen test (Supplemental Table 1). Only 2 of these patients (13%) had a personal history of an atopic disorder (both with allergic rhinitis). Conversely, 5 of 16 (31%) with a negative Phadiatop test had a history of atopy (4 patients with allergic rhinitis, one with childhood atopic dermatitis). These differences were not statistically significant (P= 0.394) Similar results were found for patients with LyP-A and LyP-C. These findings suggest that the prevalence of serologic atopy is higher than the patients’ personal recall of atopic conditions for this cohort of patients.

The lack of a correlation between patients’ history and serologic evidence of atopy for this subset of patients prompted an investigation of an additional 105 patients with CD30CLPD who were not studied for specific aero-IgEs (Supplemental Table 2). A personal history of an atopic condition was documented in the medical records of 41 (39.0%) of patients overall. For 82 patients diagnosed with LyP-A and LyP-C, an atopic condition was recorded for 35 (43%) patients which was higher than the 21% prevalence for the current cohort of LyP patients (P= 0.069). The difference was apparent for patients with LyP-A (40% vs. 11%, P= 0.024), but not LyP-C (44% vs. 50%, P= 1.0). Furthermore, the supplemental patients’ personal history of an atopic condition was associated with increased levels of IgE-t for patients with LyP-A, but not LyP-C (Supplemental Table 3). As previously shown, (3) IgE-t was also increased for LyP-A patients with a history of penicillin allergy compared to patients without penicillin allergy. These observations suggest that a link may exist between patient’s atopic diathesis and the development of LyP-A.

**Table 2.** Relationship between presence or absence of airborne or staphylococcal superantigen specific IgE and total serum IgE for categories of primary cutaneous CD30+ lymphoproliferative disease.

| Disease Category | Aero-IgE (kUa/L) | SSAg-IgE (kUa/L) | No. | IgE-t Median (range) | IgE-t GM (95% CI) |
| --- | --- | --- | --- | --- | --- |
| CD30CLPD | < 0.35 | < 0.35 | 12 | 28.8 (4.3-420) | 29.9 (12.5-71.3) |
|  | < 0.35 | ≥ 0.35 | 4 | 587 (70.8-927) | 354 (50.3-2485) |
|  | ≥ 0.35 | < 0.35 | 11 | 47.5 (20.7-598) | 66.3 (32.8-134) |
|  | ≥ 0.35 | ≥ 0.35 | 4 | 180 (33.4-491) | 145 (22.6-924) |
|  | < 0.35 | Any | 16 | 53.5 (4.3-927) | 55.4 (22.4-137) |
|  | Any | < 0.35 | 23 | 44.1 (4.3-598) | 43.7 (24.3-75.6) |
| LyP-A+C | < 0.35 | < 0.35 | 10 | 39.7 (6.6-420) | 40.3 (16.1-101) |
|  | < 0.35 | ≥ 0.35 | 4 | 587 (70.8-927) | 354 (50.3-2485) |
|  | ≥ 0.35 | < 0.35 | 11 | 47.5 (20.7-598) | 66.3 (32.8-134) |
|  | ≥ 0.35 | ≥ 0.35 | 3 | 256 (33.4-491) | 161.3 (5.0-5253) |
|  | < 0.35 | Any | 14 | 66.8 (6.6-927) | 75.0 (29.9-188) |
|  | Any | < 0.35 | 21 | 46.7 (6.6-598) | 52.3 (30.8-88.8) |
| LyP-A | < 0.35 | < 0.35 | 6 | 28.8 (6.6-420) | 40.9 (7.7-219) |
|  | < 0.35 | ≥ 0.35 | 3 | 261 (70.8-927) | 258 (10.6-6291) |
|  | ≥ 0.35 | < 0.35 | 8 | 50.5 (20.7-598) | 74.1 (26.5-207) |
|  | ≥ 0.35 | ≥ 0.35 | 2 | 262 (33.4-491) | 128 |
|  | < 0.35 | Any | 9 | 70.8 (6.6-927) | 75.6 (20.6-277) |
|  | Any | < 0.35 | 14 | 36.8 (6.6-598) |  |
| LyP-C | < 0.35 | < 0.35 | 4 | 53.5 (11.7-74) | 39.4 (10.4-150) |
|  | < 0.35 | ≥ 0.35 | 1 | 912 | ---- |
|  | ≥ 0.35 | < 0.35 | 3 | 47.5 (46.7-53.7) | 49.2 (40.7-59.5) |
|  | ≥ 0.35 | ≥ 0.35 | 1 | 128 | ---- |
|  | < 0.35 | Any | 5 | 62.8 (11.7-912) | 73.9 (20.6-277) |
|  | Any | < 0.35 | 7 | 47.5 (11.7-74.4) | 1.3 (24.7-75.9) |
| Control | < 0.35 | < 0.35 | 49 | 13.5 (2.0-110) | 12.5 (9.0-17.3) |
|  | < 0.35 | ≥ 0.35 | 3 | 132 (56.0-187) | 111 (23.9-520) |
|  | < 0.35 | Any | 52 | 14.8 (2.0-187) | 14.1 (10.1-19.9) |

Abbreviations: CD30CLPD, primary cutaneous CD30+ lymphoproliferative disease; LyP, lymphomatoid papulosis; No., number patients in cohort; IgE-t, total serum IgE (kU/L); GM, geometric mean and 95% confidence interval.

**Table 3.**
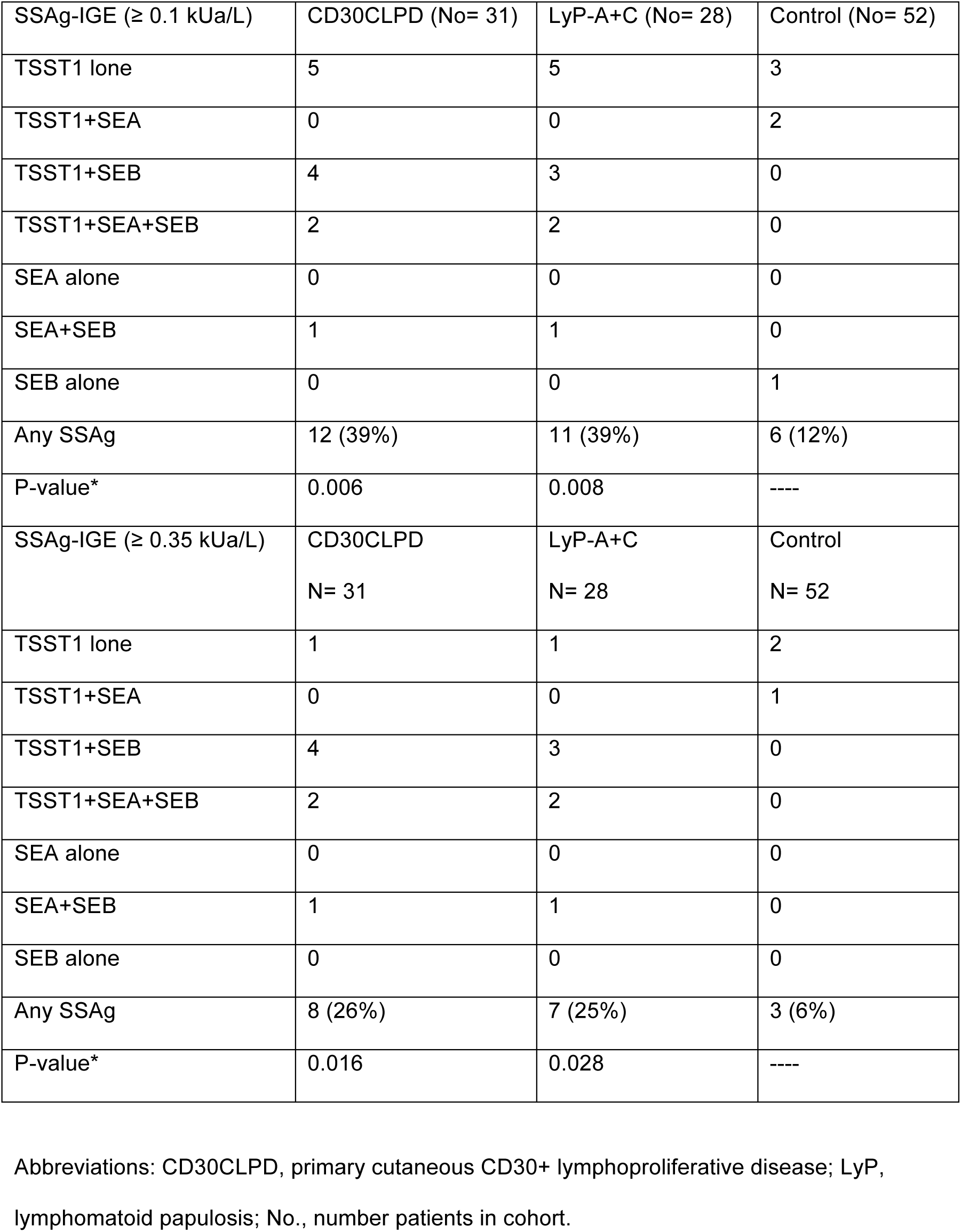
Distribution of positive Staphylococcal superantigen-specific IgE at two detection thresholds according to disease categories.

### Serologic evidence of atopy and IgE-t levels

Sixteen of 28 (57%) patients with LyP had no serologic evidence of atopy (aero-IgE < 0.35 kUa/L). The IgE-t value for these patients (median, 53.5 kU/L, range, 4.3-927 kU/L; GM, 55.4 kU/L (CI, 22.4-137 kU/L) was significantly higher than the control group (KW, P= 0.001; t-test, P= 0.002; Table 2). This finding suggests that some factor other than the patients’ atopic diathesis might be influencing IgE production.

Comparison IgE-t values for subgroups with < 0.35 kUa/L aero-IgE and < 0.35 kUa/L SSAg-IgE: IgE-t for the 3 CD30CLPD categories (KW, P= 149 and ANOVA, P= 0.257) IgE-t for LyP-A versus LyP-C subgroups (KW, P= 0.831 and t-test, P= 0.963)

Comparison IgE-t values for diagnostic categories with < 0.35 kUa/L aero-IgE and < 0.35 kUa/L: SSAg-IgE versus controls: KW, P= 0.065; ANOVA, P= 0.033

All CD30CLPD versus control: KW, P= 0.065; t-test, P= 0.059

All LyP versus control: KW, P= 0.017; t-test, P= 0.020

LyP-A only versus control: KW, P= 0.108; t-test, P= 0.131

LyP-C only versus control: KW, P= 0.047; t-test, P= 0.063

pcALCL only versus control: KW, P= 0.369; t-test, P= 0.373

### Relationship between CD30+ cells in skin lesions and IgE-t

In our previous study, (3) IgE-t GM values progressively increased for cases classified from LyP- B to pcALCL; however, the differences were not statistically significant. Given that the number of atypical CD30+ cells in the dermis generally increases from LyP-B to LyP-A to LyP-C to pcALCL, this observation suggested that CD30+ cells in skin lesions might be mediating IgE-t production. Indeed, studies in the Kadin laboratory have shown that CD30+ large atypical cells of LyP-A produce IL-13 which is an IgE-enhancing cytokine (Fig. 1). However, for the current series, IgE-t levels were about the same for LyP-A (GM, 79.2 kU/L) and LyP-C (GM, 74.1 kU/L) and lower for 3 cases of pcALCL (GM, 16.7 kU/L; Table 1).

**Fig. 1.**
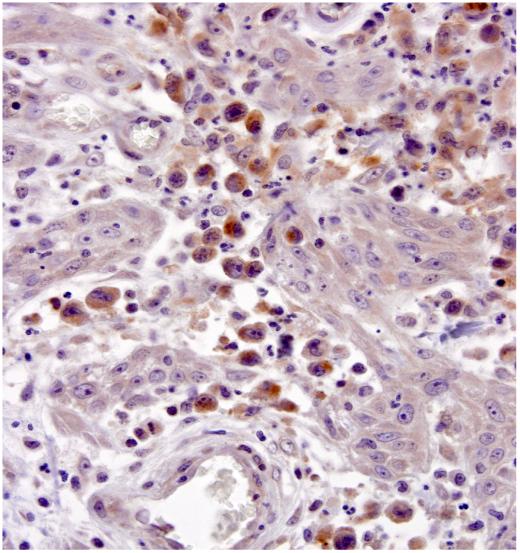
Large anaplastic cells of LyP type A express the cytokine IL-13 (brown stain). Keratinocytes and granulocytes are unstained. Immunohistochemistry was performed on formalin-fixed paraffin-embedded tissue with a murine anti-human IL-13 monoclonal antibody (1mg/ml) from Abcam (Cambridge, UK).

We therefore re-evaluated the relationship between diagnosis and IgE-t in 105 additional patients with CD30CLPD and, for comparison, 16 patients with pityriasis lichenoides, a closely related cutaneous lymphoproliferative disorder (Fig 2; Supplemental Table 4). The GMs for previously measured IgE-t (restudied cases excluded) again increased from LyP-B to LyP-C, but then decreased for patients with pcALCL. However, these differences again were not statistically significant (KW, P= 0.570; ANOVA, P= 0.474). Of interest, IgE-t values for patients with LyP-D, which expresses a CD8+CD30+ phenotype, were quite low (GM, 17.3 kU/L) and IgE-t values for pityriasis lichenoides (GM, 43.6 kU/L) were intermediate between LyP-B and LyP-A.

**Fig 2.**
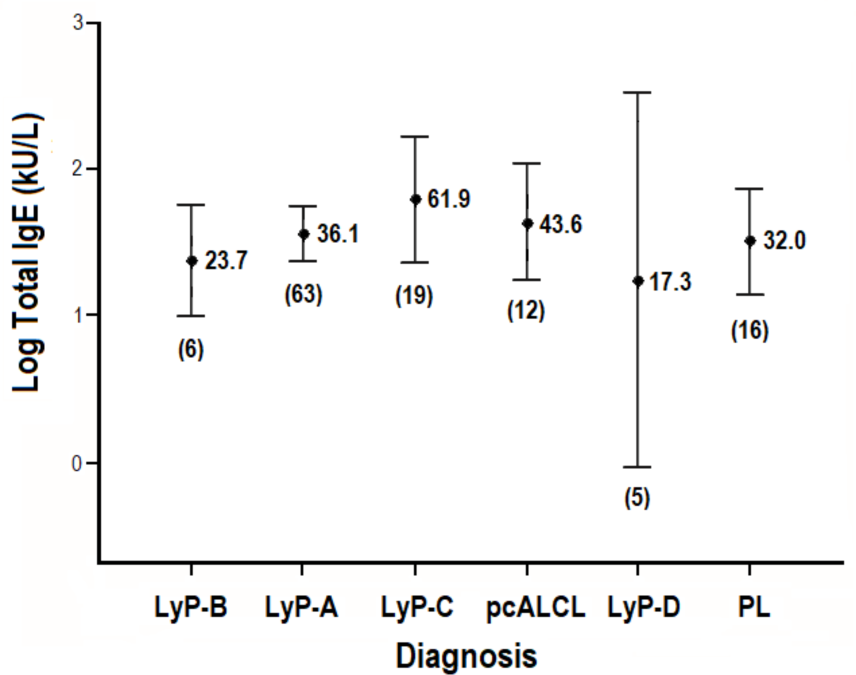
Total serum IgE values for supplemental patients with primary cutaneous CD30+ lymphoproliferative disease or pityriasis lichenoides (PL). The geometric mean (GM) and 95% confidence interval are shown for each diagnostic category. Number of patients in parentheses.

In addition, the estimated number of atypical CD30+ cells in the dermal infiltrate of skin specimens taken at the time of blood sampling was correlated to IgE-t levels. No correlation was found among categorized groups and IgE-t values (Supplemental Table 5). Moreover, no correlation was found between the level of soluble CD30 (sCD30) in the blood and IgE-t for all previously studied CD30CLPD patients (n= 76, rho, 0.149, P= 0.198; Fig 3) and LyP-A only (n= 44, rho= 0.106, P= 0.492).

**Fig 3.**
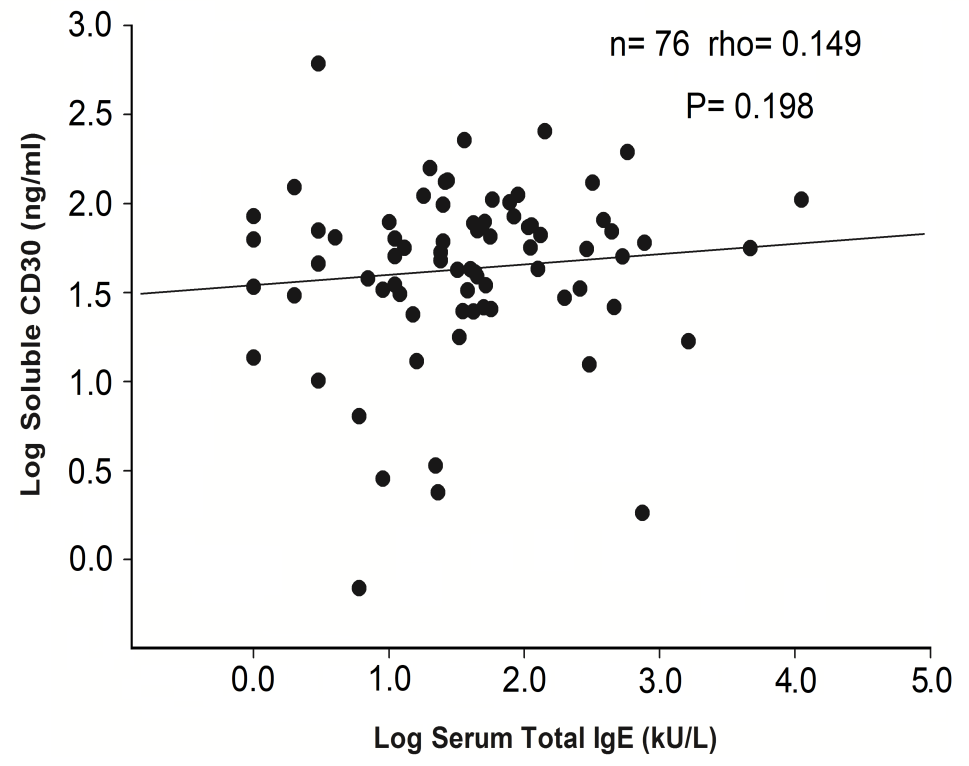
Correlation between IgE-t level and soluble CD30 level in the blood of patients with primary cutaneous CD30+ lymphoproliferative disease.

### Prevalence of Staphylococcal superantigen-specific IgE

Measurable IgE reactivity (≥ 0.1 kUa/L) against at least one SSAg was present in 12/31 (39%) specimens of CD30CLPD compared to 6/52 (12%) non-atopic control specimens (P= 0.006; Table 3). At the concentration used to define serologic atopy with aero-IgEs (≥ 0.35 kUa/L), 8/31 (26%), samples of CD30CLPD and 7/28 (25%) samples of LyP remained positive compared to 3/52 (6%) control specimens (P= 0.016 and P= 0.028, respectively). At this threshold, TSST1-IgE was detected in 7 (23%) specimens of CD30CLPD, often together with SEB-IgE (6 specimens). SEA-IgE ≥ 0.35 kUa/L was not detected. For control specimens, TSST1-IgE exceeded the 0.35 kUa/L threshold in 3 specimens (0.43, 0.47 and 0.93 kUa/L). The difference in frequency for TSST1-IgE alone remained significantly higher for all CD30CLPD samples (P= 0.002) and marginal for the LyP subset (P= 0.059).

### Relationship between SSAg-IgE and CD30+ Cells in skin specimens

The possibility that SSAgs might play a role in the proliferation of CD30+ cells of LyP was investigated by correlating the presence of SSAg-IgE with type of CD30CLPD and number of CD30+ cells in the skin specimen obtained at the time of blood sampling. Samples with SSAg-IgE values ≥ 0.1 kUa/L or ≥ 0.35 kUa/L did not occur more often for patients with higher numbers of CD30+ cells in the skin versus lower numbers, i.e., LyP-C/pcALCL vs. LyP-A or CD30+ dermal cells ≥ 20% vs. < 20%; Supplemental Table 6).

### Relationship between SSAg-IgE and serologic atopy

The relationship between serologic atopy, SSAg-IgE and IgE-t for each disease category is shown in Table 2. To minimize the influence of atopic diathesis and SSAgs on IgE-t levels, IgE-t levels of patients without a positive test for either aero-IgE and SSAg-IgE (< 0.35 kUa/L) were compared against non-atopic controls (Table 2). Although the numbers were small, IgE-t levels for patients with LyP were significantly higher than controls. No difference in IgE-t levels was apparent among the diagnostic categories, In addition, no correlation was found for IgE-t for non-atopic LyP patients categorized according to CD30 expression levels in the skin (data not shown). These observations suggest that additional factor or factors apart from the patients’atopic diathesis and perhaps presence of CD30+ cells are influencing IgE-t levels.

Table 2 also indicates that patients with CD30CLPD who are sero-negative for aero-IgEs but positive for SSAg-IgE have higher IgE-t levels compared to patients who are sero-negative to both aero-IgE and SSAg-IgE (P= 0.020). The difference was statistically significant for all LyP patients (P= 0.032), but not LyP-A alone (P= 0.122), perhaps because of the small number of patients.

Likewise, IgE-t levels were higher in patients with CD30CLPD or LyP who are sero-positive for aero-IgEs, but negative for SSAg-IgE. However, the differences were not statistically significant (P= 0.131 and P= 0.348, respectively). Consequently, IgE-t values were also high for the small number of patients who were sero-positive for both aero-IgE and SSAg-IgE.

### Systemic corticosteroid usage and IgE-t

Systemic corticosteroids might suppress production of IgE. However, IgE-t levels were significantly higher for 5 LyP-A patients who were taking systemic corticosteroids (10 to 60 mg prednisone per day) at the time of study (Supplemental Table 7). One patient had corticosteroid responsive eczematous dermatitis co-existing with LyP-A. We attribute the positive correlation between systemic corticosteroid usage and IgE-t to disease severity rather than an IgE- enhancing effect by corticosteroids.

### Smoking and IgE-t

Some studies have shown increased IgE-t in people who smoke. (7) However, in accordance with our previous study, IgE-t was not increased in patients with CD30CLPD (3)(Supplemental Table 8).

## Discussion

In this study, 50% of patients with LyP types A and C had serologic evidence of atopy (positive Phadiatop multi-allergen screen), and IgE-t was increased compared to non-atopic controls. As in our previous study, (3) no correlation was apparent between serologic atopy and the patients’ history of an atopic condition for this small cohort of patients. Nevertheless, a review of medical records of an additional 105 patients with CD30CLPD showed that IgE-t levels were increased in patients with a personal atopic history compared to patients without clinical atopy. This association was statistically significant only for LyP-A, and not for LyP-C, pcALCL or pityriasis lichenoides.

These results suggest that a pathogenic link may exist between an atopic diathesis and development of LyP-A and perhaps LyP-C. Accordingly, Nijsten noted an atopic condition in 18 of 35 (51%) cases of childhood LyP (atopic dermatitis in 2 patients, allergic rhinitis in 12 patients, allergic asthma in 3 patients, and allergic rhinitis plus asthma in 1 patient), and Miquel reported 7 of 25 (28%) children with LyP had atopic dermatitis. (10, 11) In addition, Fletcher and Laube each reported an adult patient with LyP-A and active atopic dermatitis, (12) (13) and in our series, we encountered 2 adults with long-standing atopic dermatitis and LyP (one LyP-A, one LyP-C) and several other patients with non-specific eczema. Although not observed by us, several cases of pcALCL occurring in patients with atopic dermatitis have been reported, often in the context of cyclosporine therapy given for the atopic dermatitis. (14) (15)

In agreement with our previous study, (3) IgE-t GM values of supplemental patients progressively increased from LyP-B to LyP-A to LyP-C, and with increasing categories of CD30+ dermal cells below 50% in the skin specimen obtained at the time of blood sampling. This observation suggests that the atypical CD30+ cells might be influencing IgE production, and studies in the Kadin laboratory have shown that CD30+ large atypical cells of LyP-A are capable of producing IL-13 which is an IgE-enhancing cytokine (Fig. 1). However, if CD30+ cells directly influence IgE-t production, then the decrease in IgE-t for patients diagnosed to have pcALCL compared to LyP-C requires explanation. One possibility is that the total biomass of CD30+ cells in pcALCL may be less than LyP-C. This might occur because pcALCL is characterized by fewer skin lesions than those of patients with LyP-C, and the infiltrate of some pcALCL lesions may have fewer than 50% CD30+ cells, thereby resembling LyP-A.(2) Nevertheless, the lack of a correlation between IgE-t number of CD30+ cells in skin specimens (Supplemental Table 5) and sCD30 blood levels (Fig 3) suggest that CD30+ cells per se are not the major factor mediating IgE-t levels. What factors other than number of CD30+ cells might account for the increased IgE in LyP? Patients’ age and gender, smoking history and use of systemic corticosteroids are seemingly not important.

In addition to atopic diathesis, our study indicates that SSAgs might be involved in stimulating IgE production given that IgE-t levels were higher for non-atopic patients with SSAg-IgE levels ≥ 0.35 kUa/L than patients with < 0.35 kU/L (Table 2). The detection of SSAg-IgEs in nearly 50% of patients with LyP indicates that exposure to bacterial superantigens, particularly TSST1, has taken place. Although overt clinical infection by S. aureus is not typical of LyP lesions, it is intriguing that some patients with LyP improve on oral antibiotics in Dr. Vonderheid’s experience. Moreover, the onset of LyP (and the closely related pityriasis lichenoides) (16) sometimes seems to be triggered by an infection.(17) (11) Given that Streptococcal superantigens have a pathogenic role in eruptive guttate psoriasis, could a similar mechanism occur in LyP?(18);(19) If so, the atypical cells of LyP might have a skewed TCR-Vβ profile, e.g., increased TCR-Vβ2 expression which corresponds to stimulation by TSST1 superantigen. Thus far, data to support or refute this hypothesis are limited.(20)

An alternative hypothesis that provides linkage with atopy might also be considered. If CD30+ cells are not the primary source of IgE-enhancing cytokines, then other cells in the inflammatory infiltrate of LyP lesions might be important. For example, it seems possible that S. aureus at ulcerative LyP lesion sites might activate skin resident type 2 innate lymphoid cells (ILC2) which then secrete IL-5 and IL-13, express CD40L (CD154) and interact with CD40+ B-cells to increase IgE including SSAg-IgE. To account for the high IgE-t levels in LyP types A and perhaps C, LyP patients with an atopic diathesis have hyper-responsive and/or increased numbers of ILC2 cells in the skin compared to non-atopic individuals as has been reported for atopic dermatitis. (21) (22) (23) The relationship between atopic diathesis and ILC2 would also explain why IgE-t is often increased in a variety of other inflammatory skin diseases.(3) Further studies are required to test this hypothesis.

LyP is clinically benign but histologically malignant.(24) This paradox may be in part explained by the high rate of apoptosis of the CD30+ anaplastic cells in regressing LyP skin lesions. (25) In this regard it is interesting to note that bacterial SSAgs have been shown to induce apoptosis of neoplastic T cells in mice leading to improved survival of lymphoma bearing mice. (26) To test this hypothesis in LyP would require isolation of CD30+ cells from skin lesions and in vitro exposure to cognate Vβ superantigens. This research could be aided by identification of IgE specific antibodies to SSAgs in the current study.

A limitation of this study is the relatively small number of patients for whom sera and corresponding clinical histories were available, largely due to the rarity of CD30CLPD. A second limitation is analysis of only 3 SSAgs as a larger number SSAgs might disclose further evidence of IgE specific antibodies. Finally, we analyzed only IgE (not IgG) specific antibodies.

In summary, patients with the most common LyP types A and C have serologic evidence of atopy against common airborne antigens and SSAgs when compared to control adult subjects who had rhino-sinusitis and a negative Phadiatop test for aero-IgEs. Serologic evidence of atopy exceeded that determined by LyP patients’ personal history. The results suggest that an atopic diathesis and perhaps SSAg may contribute to the pathogenesis of LyP

## Data Availability

Article DOI: 10.1111/1346-8138.15059

